# Detection of pancreatic beta cell mass in vivo in humans: studies in individuals with long-standing type 1 diabetes and in individuals with obesity

**DOI:** 10.64898/2026.03.12.26348138

**Authors:** Alessandra Dei Cas, Valentina Spigoni, Raffaella Aldigeri, Federica Fantuzzi, Gloria Cinquegrani, Elisabetta Giordano, Roberta Eufrasia Ledda, Valentina Casale, Silvia Migliari, Maura Scarlattei, Livia Ruffini, Riccardo C. Bonadonna

## Abstract

**Background:** PET-CT scans of radioactive exendin-4, a ligand of the GLP-1 receptor, are claimed to provide a biomarker of pancreatic beta cell mass (BCM), although the GLP-1 receptor is expressed also in the exocrine pancreas (PX). Parotid glands may be a reference tissue for GLP-1 receptor expression in exocrine cells of the GI system. Our aims were 1. To assess biomarker(s) of BCM derived from ^68^Ga-NODAGA-exendin-4 PET-CT scans in participants with long-standing type 1 diabetes (T1DM) or in subjects with obesity (OBESE); 2. To investigate the relationship between biomarker(s) of BCM and a biomarker of beta cell functional mass (BCFxM) in OBESE.

**Methods:** T1DM (n=8, Age: 50.4±3.8 yrs; T1DM duration: 34.2±3.0 yrs; BMI: 26.6±1.1 kg/m^2^; HbA1c: 7.5±0.36%) and OBESE (n=9; Age:48.2±2.2 yrs; BMI: 37.4±1.1 kg/m^2^; HbA1c: 5.4±0.17%) underwent two studies: 1) ^68^Ga-NODAGA-exendin-4 PET-CT scan of both PX and parotid glands 45’-60’ after i.v. injection and with CT-assessment of PX volume to compute biomarkers of BCM based on SUV (BCM_SUV_) or clearance (CLEAR; BCM_CLEAR_); 2) Mixed meal test (MMT), with measurement of plasma glucose, C-peptide, GLP-1 and GIP curves to assess BCFxM with state-of-art mathematical modeling.

**Results:** The C-peptide response to the MMT in T1DM participants was absent or negligible, whereas the OBESE displayed a robust BCFxM. The PX volume was smaller in T1DM than in OBESE (51.7±6.6 vs 92.9±10.9 cc; p=0.007). The biomarkers of BCM, as assessed by ^68^Ga-NODAGA-exendin-4 SUV or CLEAR, were 6.6-fold (p=0.003) and 5.0-fold (p=0.002) lower, respectively, in T1DM than in OBESE. BCFxM was correlated in OBESE to both biomarkers of BCM (r=0.91 p<0.001, and r=0.82 p=0.006, respectively).

**Conclusion/interpretation:** ^68^Ga-NODAGA-exendin-4 derived biomarkers of BCM can discriminate T1DM from OBESE. In OBESE ^68^Ga-NODAGA-exendin-4 derived BCM appears to be a pivotal determinant of the beta cell response to MMT and may be valuable to compare and monitor BCM both in research and in clinical settings.

**Research in context:** *What is already known about this subject?:*

- Changes in pancreatic beta cell functional mass are at the heart of alterations in glucose regulation, including diabetes mellitus. Beta cell functional mass can be assessed by mathematical modeling of the in vivo beta cell response to intravenous or oral challenges.
- Beta cell functional mass is the product of beta cell mass times beta cell function per mass unit, i.e. the result of two distinct entities, mass and function. No in vivo methods can dissect out beta cell mass and function.
- Pancreatic ^68^Ga-exendin-4 uptake, as measured by PET-CT, has been proposed as a non-invasive biomarker of beta cell mass. However, the ratio of 3.6:1 between endocrine and exocrine pancreas ^68^Ga-exendin 4 uptake suggests that there is room for improvement.

*What are the key questions?:*

- Does an improved ^68^Ga-exendin4 method provide a better separation between participants with type 1 diabetes and expected zero/nil beta cell mass vs people with nondiabetic obesity?
- What is the role of beta cell mass in determining beta cell functional mass in people living with obesity?

*What are the new findings?:*

- The improvement in the quantitation of beta cell ^68^Ga-exendin-4 binding to beta cells resulted in a clearcut separation of participants with type 1 diabetes and expected zero/nil beta cell mass from people living with obesity
- In people living with obesity, beta cell mass, as assessed by ^68^Ga-exendin-4 PET-CT scan, is a pivotal determinant of beta cell functional mass, as assessed by mathematical modeling of a frequently sampled mixed meal test

*How might this impact on clinical practice in the foreseeable future?:*

- This method has the potential to track changes in beta cell mass both between-subjects and within-subjects over time
- Natural history of glucose (in)tolerance and the impact of disease modifier candidates in diabetes mellitus can be assessed with the present method

## Introduction

Pancreatic beta cell functional mass can be defined as the product of beta cell mass (BCM) multiplied by the average beta cell function per mass unit, i.e. the compound measure of two distinct entities, mass and function per mass unit. The in vivo methods to assess “insulin secretion” or “beta cell function” in humans can provide more or less accurate estimates of beta cell functional mass, as defined above (1).

Changes in beta cell functional mass are at the heart of alterations in glucose regulation. Both in type 1 (2) and in type 2 diabetes (3), albeit with quite different etiopathogeneses, the decline of beta cell functional mass is the pacemaker of worsening glucose regulation both before and after the diagnosis. To a considerable extent, the natural history of diabetes is the natural history of the decline of beta cell functional mass (4).

Classical methods exist to quantify beta cell functional mass in vivo, including both intravenous and oral administration of secretagogue nutrients (1). In the last two decades mathematical modeling of glucose and C-peptide curves after oral or intravenous challenges has become a reference approach to assess beta cell response in vivo in humans (1). Nevertheless, also modeling derived parameters do not overcome the intrinsic limitation of tracking only beta cell functional mass, with no possibility of dissecting out mass and function.

Several human studies have exploited either cadaveric pancreata (5)(6)(7)(8) or pancreatic tissue samples obtained from surgical interventions (9–11) to explore the role that BCM plays in glucose regulation. These contributions have illuminated our understanding of the pathogenesis of both type 2 and type 1 diabetes. Nevertheless, a number of issues remain unsolved: i. autoptic studies usually have no assessment of beta cell functional mass in vivo (5,6,8); ii. surgical studies generally rely on measures of percent of beta cells in pancreas slices, i.e. beta cell density (9), an incomplete biomarker of BCM, which equals the product of beta cell density times pancreatic mass; iii. surgical removal of fractions of the whole pancreas are the best available, but largely imperfect, model of the effects subsequent to partial beta cell loss in humans (10). The exact role, therefore, played by BCM in determining beta cell functional mass still is unclear.

If a noninvasive in vivo method were to provide a reasonable biomarker of BCM, its combination with one of the recognized methods to assess beta cell functional mass could enable also to assess the relationship between mass and function in humans, the latter being computable as the ratio of beta cell functional mass divided by BCM. Such a tool would be of great help in assessing better the natural history of the different forms of diabetes mellitus and in monitoring the effects of different anti-diabetes therapies, especially regarding their potential as disease modifiers.

Several noninvasive methods have been proposed to quantify separately BCM in humans. Most of them have relied on the intravenous injection of radioactive ligands putatively specific for pancreatic beta cells (12). For instance, a VMAT2 ligand was extensively studied for its potential to be a biomarker of pancreatic beta cell mass (13). However, in humans VMAT2 is expressed also in islet cells other than beta cells (14). In line with this premise, the VAMT2 ligand ^11^C-dihydrotetrabenazine has shown a less than optimal potential to discriminate between healthy controls and people with type 1 diabetes (15). Similar limitations were reported with ^11^C-hydroxytryptophan, which is a preferential ligand of beta cells, but it also binds to other islet endocrine cells (16).

Pancreatic uptake of radiolabeled exendin-4, a ligand of glucagon-like peptide-1 receptor, can be detected by positron emission tomography (PET) (17). It has been claimed to provide a non-invasive biomarker of BCM (18), but it still is unclear whether it can provide, for instance, a clearcut separation between participants with type 1 diabetes, i.e. with expected close to zero/nil BCM, and people with expected robust BCM. The use of radiolabeled exendin-4 in humans to generate a BCM biomarker has been criticized, because, as recently confirmed, in humans the median pancreas endocrine /exocrine ratio of radioactive exendin-4 uptake is 3.6:1 (19), with a wide spread, whereas in rodents it is about 45:1. Thus, in man pancreatic uptake of exendin-4 might provide a quite spurious signal.

Nevertheless, among type 1 diabetes participants those with better and less variable 24h glucose profiles display higher pancreatic ^68^Ga-NODAGA-exendin-4 uptake, as detected by PET-CT, in agreement with the expectation that their residual BCM should be higher (20). Furthermore, in people with pancreatic islet transplantation there is a detectable uptake of ^68^Ga-NODAGA -exendin-4 in the liver, raising the possibility that PET-CT scan of ^68^Ga-NODAGA-exendin-4 could be used to monitor the fate of transplanted islets (21). Finally, PET-CT scans with ^68^Ga-exendin currently are under study to predict the response to dual agonists of entero-hormones in individuals with type 2 diabetes (22).

In spite of these remarkable achievements, we thought that there still is room for significant improvement of the ^68^Ga- NODAGA-exendin-4, for at least two reasons. First, most of previous papers have reported pancreatic standardized uptake values (SUVs) of the radioligand under study as the biomarker of BCM, failing to recognize that SUV tracks density of beta cells per volume unit, but an estimate of BCM would require also the measurement of pancreatic volume. Secondly, as alluded above, pancreatic exocrine cells express GLP-1 receptor, thereby blurring the specificity of exendin-4 binding for the beta cells. This potential shortcoming could be overcome with the use of a reference region to be used as a correction for the ^68^Ga-NODAGA-exendin-4 uptake to be ascribed to pancreatic exocrine cells. Candidates for this task are the salivary glands, which, beyond their histological similarity to the pancreatic exocrine tissue, also exhibit a significant expression of the GLP-1 receptor (23)(24).

The present investigation, therefore, was undertaken to assess the performance of an improved ^68^Ga-exendin-4 PET-CT method in separating people with long-standing type 1 diabetes, supposedly with (almost) nil beta cell mass, from people living with obesity, supposedly with large beta cell mass. An additional goal of this study was to explore the relationship, if any, between the ^68^Ga-exendin-4 derived estimate of BCM and the beta cell functional mass, as assessed by a reference method.

## Research Design and Methods

### Subjects

Participants with long-standing type 1 diabetes mellitus were recruited in the Diabetes Outpatient Clinic of the Division of Endocrinology and Metabolic Diseases, Azienda Ospedaliero-Universitaria di Parma (Parma, Italy). Participants with nondiabetic obesity were recruited in the Outpatient Clinic of the Unit of Alimentary and Metabolic Sciences, Azienda Ospedaliero-Universitaria di Parma (Parma, Italy). Participants with diabetes had to have diabetes for at least 15 years as per experimental design.

All individuals were studied twice in random order, after an overnight fast. On one occasion they underwent a mixed meal tolerance test (see below). On the other occasion they underwent a ^68^Ga-NODAGA-exendin-4 PET-CT scan (see below). The study (ClinicalTrials.gov number: NCT 05662189) was reviewed and approved by the Area Vasta Emilia Nord Ethical Committee (approval prot. n. 41494 of 14^th^ October 2021) and all participants were included in the study after signing an informed written consent.

Eight people with long-standing type 1 diabetes and ten subjects with obesity agreed to participate and completed both sessions of this study. After revising the PET-CT data of one person with obesity, it was found that his pancreatic volume was 287 cm^3^, i.e. 6 standard deviations above the mean value of the other subjects with obesity and 4 standard deviations above the subject with the second highest value of pancreatic volume. Since the pancreatic volume is a key measure of this research, it was decided to include no data of this outlier individual in the present report. Thus, all data of the group formed by people with obesity are derived from nine subjects.

### Mixed-meal tolerance test

A standardized 5 h mixed meal test (MMT) was performed in the morning after an overnight fast. The MMT provided 1.628 MJ (386 Kcal; 52% CHO, 17% proteins, 31% fats). The time taken by the subjects to ingest the meal was recorded. Time 0’ was the moment in which the subjects started to ingest the MMT. Plasma glucose, C-peptide, total GLP-1 and total GIP were measured at time - 20,-10, 0, 10, 20’, 30, 45, 60, 90, 120, 150, 180, 210, 240, 270, and 300 minutes.

Glycated hemoglobin, blood count, lipid profile, liver and renal function parameters were measured by standard in-house methods.

Plasma glucose concentration (mmol/l) was measured with the YSI 2300 Stat Plus Glucose&Lactate Analyzer (YSI Inc., Yellow Springs, OH, USA).

Venous blood samples were collected for hormone determinations at each time point into pre-chilled tubes containing a protease inhibitor cocktail to prevent degradation of peptide hormones. Tubes contained 100 µM dipeptidyl peptidase-4 (DPP-4) inhibitor (Merck Millipore, Darmstadt, Germany), 2 mM Pefabloc SC Plus (Roche Diagnostics, Basel, Switzerland), 1 µM aprotinin (Sigma-Aldrich, St. Louis, MO, USA), and 10% disodium EDTA.

Immediately after collection, samples were kept on ice and centrifuged at 3500 × g for 10 minutes at 4 °C. Plasma was separated and stored at −80 °C until analysis.

Plasma concentrations of C-peptide in people with obesity were measured using a commercially available ELISA kits from Mercodia (Uppsala, Sweden). In participants with long-standing type 1 diabetes, C-peptide concentrations were quantified using an ultrasensitive ELISA (Mercodia, Uppsala, Sweden) to enable detection of very low circulating levels. Circulating levels of total glucagon-like peptide-1 (GLP-1) and glucose-dependent insulinotropic polypeptide (GIP) were determined using ELISA kits from Merck Millipore (Darmstadt, Germany). Internal quality control standards provided with the GLP-1 and GIP kits were included in each assay run to verify assay performance and ensure that measured concentrations fell within the expected ranges.

All assays were performed according to the manufacturers’ instructions, with assay sensitivity and intra- and inter-assay coefficients of variation within the specified ranges. Samples were analyzed in duplicate, and samples from the same participant were analyzed within the same assay run to minimize inter-assay variability. Absorbance for Mercodia assays was measured at 450 nm, whereas for GLP-1 and GIP assays absorbance was measured at 450 nm with wavelength correction at 590 nm using a Multiskan™ FC microplate reader (Thermo Fisher Scientific, Waltham, MA, USA).

### Assessment of pancreatic beta cell functional mass

Beta-cell functional mass was estimated by mathematical modeling, as previously described (25)(26), modified to account for the additional contribution of incretins (GLP-1 and GIP). The mathematical model was implemented in the SAAM 2.2 software (27). Briefly, the beta cell response to the meal is described as the sum of three components: the first one is related to the rate of increase of plasma glucose (DC: derivative control), the second one related to glucose concentration *per se* (PC: proportional control, otherwise denominated glucose sensitivity of the beta cell). A third component accounted for a further contribution to total insulin secretion rate ascribable to GLP-1/GIP, when detectable. Please note that this third component does not measure the total incretin effect, most of which is already embedded in PC, which in turn accounts for most of total insulin secretion rate and, therefore, has been selected as the biomarker of beta cell functional mass in this work. PC, or beta cell glucose sensitivity, in this paper is presented as the slope of the curve relating plasma glucose concentration to the response in insulin secretion rate during the meal. Please, see the Supplementary Material for further details.

### Radiopharmaceutical Preparation

In the Radiopharmacy Laboratory of the Nuclear Medicine Division, [^68^Ga]Ga-NODAGA-exendin-4 was synthesized following the radiolabelling method previously described in detail (28) (29) using a 68Ge/68Ga generator (GalliaPharm®, Eckert & Ziegler, Berlin, Germany; nominal activity 1850 MBq), certified for GMP compliance and aligned with the European Pharmacopoeia monograph [Gallium (CCGa) Chloride solution for radiolabelling, Monograph 2464], and an automated synthesis module (Scintomics GRP® module, Fürstenfeldbruck, Germany) equipped with a single-use disposable cassette (SC-01, ABX, Radeberg, Germany), both placed in a GMP-compliant, ISO Class 5 (Grade A) hot cell (NMC gallium-68, Tema Sinergie, Faenza, Italy), enabling aseptic production conditions.

The disposable single-use cassette was equipped with a strong cationic exchange (SCX) cartridge for the cationic purification of the eluate 68 GaCl 3.

The peptide precursor NODAGA-exendin-4 was purchased as lyophilized GMP-grade powder from piCHEM (Forschungs- und Entwicklungs Grambach, Austria).

All reagents employed in the radiolabelling process—including sodium chloride (NaCl), ethanol, 2-[4-(2-hydroxyethyl)-1-piperazinyl]-ethanesulfonic acid (HEPES) buffer, phosphate-buffered saline (PBS), and water for injection (WFI) — were of the highest pharmaceutical purity and provided as a single-use synthesis kit (SC-102, ABX, Radeberg, Germany), ensuring batch-to-batch consistency and compliance with current Good Manufacturing Practice (cGMP) requirements.

Analytical-grade, metal-free reagents used for quality control procedures of 68Ga-NODAGA-Ex4, including trifluoroacetic acid (TFA), water, and acetonitrile for radioactivity and ultraviolet-coupled high-performance liquid chromatography (Radio-UV-HPLC), as well as ammonium acetate and methanol for radiochemical purity assessment, were purchased from Sigma-Aldrich (St. Louis, MO, USA).

Relevant quality control parameters such as efficacy of radiolabelling procedure (radiochemical yield, RY% and molar activity, Am) and quality of ^68^Ga-NODAGA-Ex4 (radiochemical purity, RCP%) were assessed for each produced batch following the analytical methods previously developed and validated (29).

## 68Ga-NODAGA-exendin-4 PET-CT Scan

All the studies were carried out in the morning after an overnight fast. The subject was positioned on the scan bed in the supine position with arms over the head. A slow i.v. intravenous injection of the PET tracer was performed immediately before image acquisition. Dynamic images were acquired over 20 minutes (1 bed) at the junction between thorax and abdomen (8 frames, 15 seconds each, over 2 minutes, 10 frames, 30 seconds each over 5 minutes and 13 frames, 60 seconds each over 13 minutes) using a 3D integrated PET/CT system (Discovery IQ, GE Healthcare®). Subsequently, a whole body scan from the orbitomeatal line to the upper abdomen (3 beds, 3 minutes each) was performed 45-60 minutes after tracer injection with the subject in the same position of dynamic images.

A low dose CT (130Kv, 40 mAs, pitch 1, collimation 16 x 1.25, 1 bed position) was used for attenuation correction and to provide an anatomical reference.

Attenuation, normalization, random, decay, and scatter corrections were performed. PET data were reconstructed with the Q Clear algorithm (a Bayesian penalized-likelihood reconstruction algorithm, GE Healthcare®, strength 350).

Low dose PET-CT images, retrieved from the institutional Picture Archiving and Communication System (PACS), were analyzed by a single operator (radiologist with more than four years of experience) using a dedicated workstation and quantitative imaging software (MM Reading, Syngo.via, Siemens Healthineers) for semi-automated segmentation and subsequent calculation of pancreatic and renal volume and density and parotid gland density.

Pancreatic and renal margins were manually delineated by the operator using regions of interest (ROIs) drawn on one out of every two CT slices, with automatic interpolation of the intermediate slices. The software then automatically generated a volume of interest (VOI, measured in cm³) and provided maximum, minimum, and mean density values, expressed in Hounsfield Units (HU). In cases of inaccurate segmentation, manual adjustment of the VOI boundaries was allowed.

Parotid gland density was evaluated by placing a fixed-size ROI of 35 cm² within both glands.

### Quantitative Analysis

Quantitative analysis of tracer uptake was performed on a workstation Advantage 4.4 (GE Healthcare, Milwaukee, WI) using spherical volumes of interest (VOIs) semi-automatically drawn on the pancreatic segments (head, body and tail) and the parotid glands Standardized uptake value (SUV) was measured as the ratio of decay-corrected activity in the VOI (KBq/mL) to the injected activity per unit body weight. The SUVmax was calculated as the hottest voxel within the VOI, while SUVmean takes in account all the voxels contained in the VOI. SUVmax and SUVmean of pancreatic segments and parotid glands were measured on static images acquired at 45-60 minutes. Uptake of the whole pancreas was obtained as the average value across head, body and tail segments. The average value between the two parotid glands for both SUVmax and SUVmean was calculated for each scan.

### Computation of Biomarkers of Beta Cell Mass

Pancreatic ^68^Ga-NODAGA-exendin-4 uptake (45’-60’ after injection) was averaged across head, body and tail.

Two slightly different methods were used to compute a biomarker of BCM. In the first method the following equation was used:

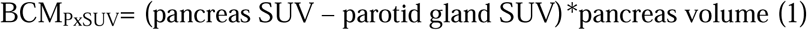

At any given time point, equation 1 computes the fraction of the exendin-4 dose taken up by the endocrine pancreas multiplied by body weight (grams). Its units are pure number.

In the second method the following equation was used:

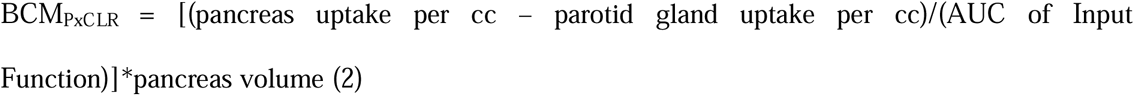

At any given time point, equation 2 computes the apparent exendin-4 clearance by the endocrine pancreas. Its units are: ml . min^-1^.

Equation 2 requires the use of the input function of ^68^Ga-NODAGA-exendin-4; hence it is experimentally more cumbersome.

### Statistical analysis

Descriptive statistics are reported as mean ± standard error of the mean (SEM) for continuous variables and frequency for categorical variables. Comparison between groups was performed by means of t-test, or generalized linear models for repeated measures, or χ2 test for categorical variables, as appropriate. All statistical analyses were carried out with the SPSS 30.0 software. Statistical significance was declared at p<0.05.

## Results

The main clinical and metabolic features of the study participants are reported in Table 1. The two groups differed as to the balance women/men, BMI, fasting plasma glucose and HbA1c.

**Table 1.** Demographic, anthropometric, clinical features and administered exendin-4 doses of the subjects participating in this research.

|  | **T1DM** | **OBESE** | **P value** |
| --- | --- | --- | --- |
| Subjects (M/F) | (8/0) | (4/5) | 0.01 |
| Age (yrs) | 50.4±3.9 | 48.2±2.2 | 0.63 |
| Diabetes Duration (yrs) | 34.2±3.0 | - |  |
| BMI (kg/m<sup>2</sup>) | 26.6±1.1 | 37.4±1.9 | <0.001 |
| Fasting Glucose (mmol/L) | 6.5±0.7 | 5.1±0.2 | <0.01 |
| HbA1c (%) | 7.5±0.36 | 5.4±0.17 | <0.001 |
| Pancreas Volume (cc) | 51.7±6.6 | 92.9±10.9 | 0.007 |
| <sup>68</sup>Ga-NODAGA-exendin-4 Dose (MBq) | 104±2.0 | 135±7.3 | 0.001 |
| Exendin-4 peptide Dose (µg) | 2.19±0.10 | 3.13±0.27 | 0.006 |

T1DM: Participants with type 1 diabetes; OBESE: Subjects with obesity

The glucose curve after the MMT in participants with type 1 diabetes peaked at 120 min and was not back to baseline at 300’ (Figure 1a). In subjects with obesity, the glucose curve after the MMT peaked at 30’ and was back to baseline by 240’ (Figure 1a). On the day of the MMT, three subjects with obesity displayed fasting plasma glucose concentrations compatible with the state of impaired fasting glucose (i.e. ≥ 5.6 mmol/L).

**Figure 1.**
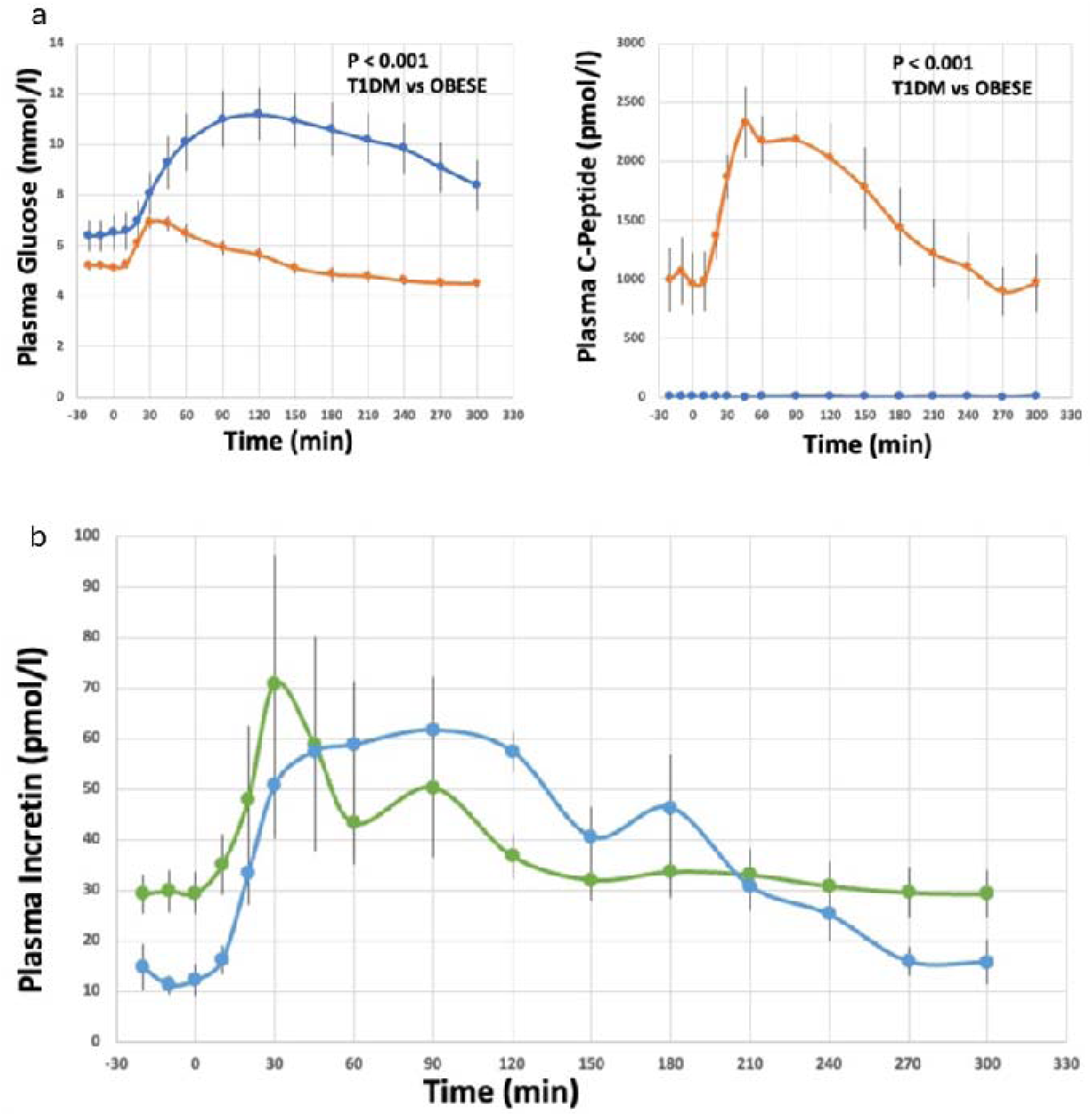
(a) Left Panel: plasma glucose levels in people with type 1 diabetes (T1DM) and in subjects with obesity (OBESE) before and after the ingestion of the mixed meal. Right Panel: plasma C-peptide levels in people with type 1 diabetes (T1DM) and in subjects with obesity (OBESE) before and after the ingestion of the mixed meal. (b) Plasma GLP-1 and GIP levels in in people with obesity before and after the ingestion of the mixed meal.

The C-peptide response to the meal was negligible or absent in all participants with type 1 diabetes, so that no modeling of beta cell function was feasible (Figure 1a). As a consequence, the estimate of their beta cell functional mass should be considered zero, or very close to zero. In the subjects with obesity, a robust C-peptide response was detected (Figure 1a).

The plasma glucose (Figure 1a), C-peptide (Figure 1a), and GLP-1 and GIP curves (Figure 1b) of the subjects with obesity were used to run the mathematical model of the response of the beta cells to the MMT. In the subjects with obesity, the proportional control of beta cell (otherwise denominated beta cell glucose sensitivity), which is the biomarker of beta cell functional mass, was 426±96 pmol.min^-1^ of insulin per mmol.l^-1^ of plasma glucose.

### PET-CT scans

The scans of one representative participant with type 1 diabetes and of one representative subject with obesity are shown in Figure 2. Injected dose of 68Ga-NODAGA-exendin-4 and peptide amount are reported in Table 1. Radiosynthesis results are reported in the Supplementary materials.

**Figure 2.**
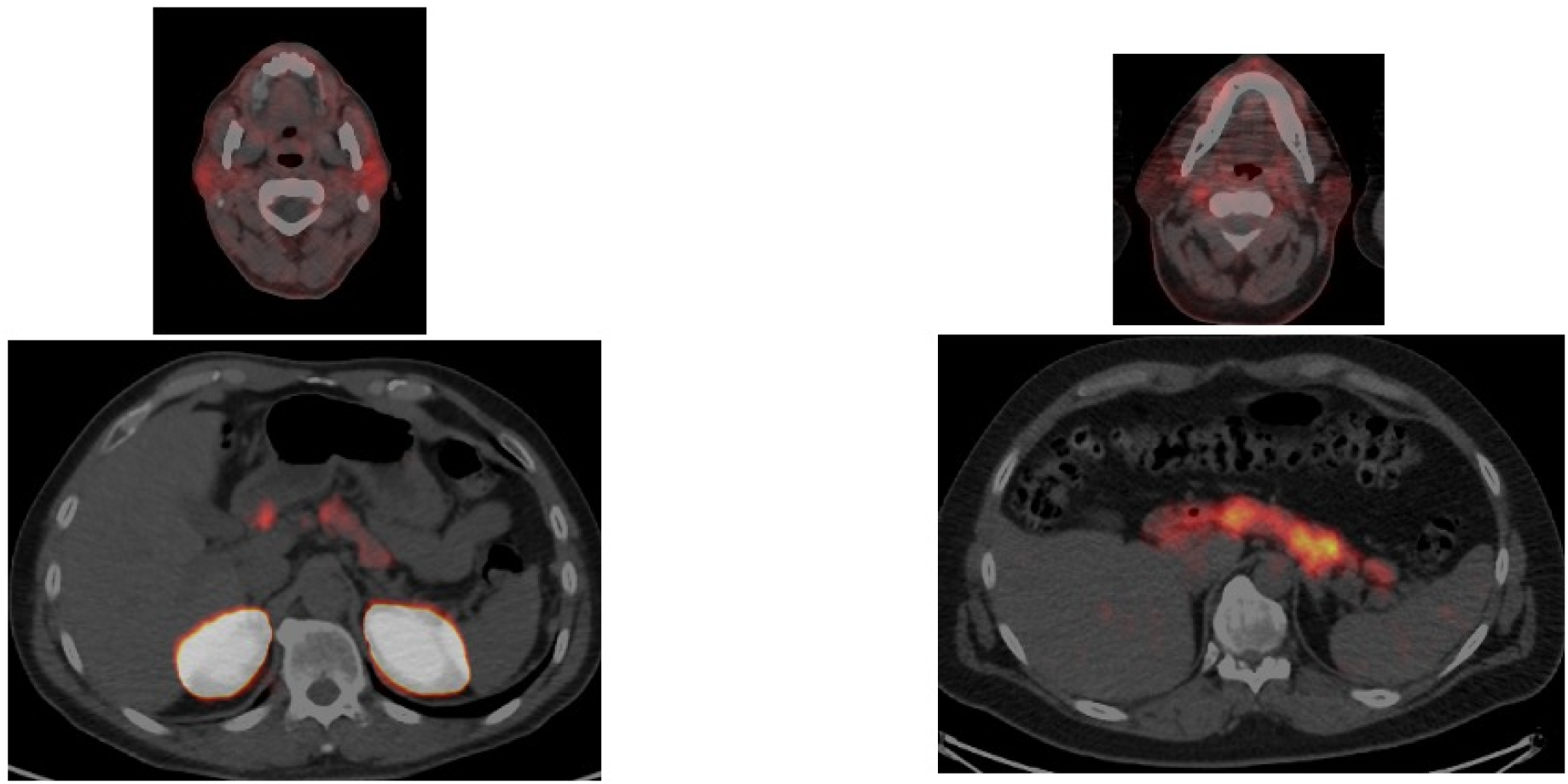
^68^Ga-NODAGA-exendin-4 PET-CT of parotid glands (upper images) and pancreas (bottom images) in one representative individual with type 1 diabetes (left) and one representative subject with obesity (right).

Analysis of the PET-CT images showed that in the pancreas both ^68^Ga-NODAGA-exendin-4 SUVmean and ^68^Ga-NODAGA-exendin-4 clearance were significantly lower in participants with type 1 diabetes than in subjects with obesity, whereas in the parotid glands both parameters were similar in the two groups (Figure 3a and 3b). When the parotid glands were used as the reference region of pancreatic ^68^Ga-NODAGA-exendin-4 uptake, the differences between people with type 1 diabetes and people with obesity in the biomarkers of pancreatic beta cell density were striking (Figure 3a and 3b). Pancreatic volume also was significantly smaller in the participants with type 1 diabetes than in the subjects with obesity (Table 1). As a result, there was an almost complete separation (Figure 3a and 3b) in both biomarkers of BCM, the one based on ^68^Ga-NODAGA-exendin-4 SUV (BCM_SUV_) and the one based on ^68^Ga-NODAGA-exendin-4 apparent clearance (BCM_CLEAR_) between the participants with type 1 diabetes (BCM_SUV_: 32.2±7.8; range: 8.5 - 71.8 --BCM_CLEAR_: 219±37.3; range: 62.1 - 357) and the subjects with obesity (BCM_SUV_: 213±47.1; range: 53.4 - 492 -- BCM_CLEAR_: 1104±218; range: 276 - 2452).

**Figure 3.**
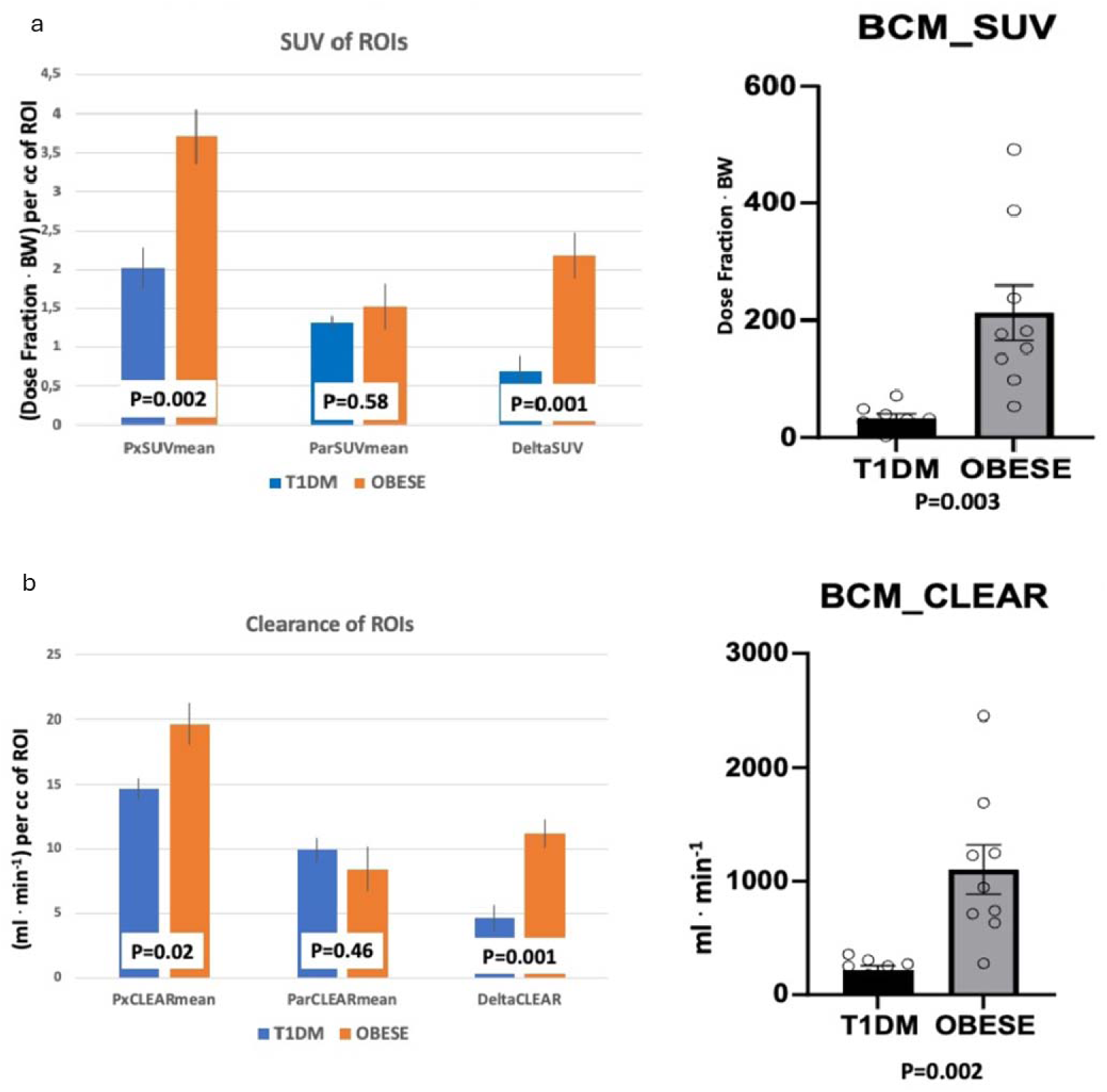
(a) 68Ga-NODAGA-exendin-4 SUV of ROIs (left panel) and biomarker of beta cell mass in the pancreas (BCM_SUV; right panel) in participants with type 1 diabetes (T1DM; histograms on left) and subjects with obesity (OBESE; histograms on right). (b) 68Ga-NODAGA-exendin-4 clearance of ROIs (left panel) and biomarker of beta cell mass computed with 68Ga-NODAGA-exendin-4 clearance (BCM_CLEAR; right panel) in participants with type 1 diabetes (T1DM; histograms on left) and subjects with obesity (OBESE; histograms on right). SUV: standardized uptake value; ROI: region of interest; BW: body weight; PxSUVmean: average SUV in the pancreas; ParSUVmean: average SUV in the parotid glands; DeltaSUV: PxSUVmean minus ParSUVmean, biomarker of beta cell density in the pancreas; PxCLEARmean: average clearance in the pancreas; ParCLEARmean: average clearance in the parotid glands; DeltaCLEAR: PxCLEARmean minus ParCLEARmean, biomarker of beta cell density in the pancreas; BCM_SUV: biomarker of beta cell mass in the pancreas computed with ^68^Ga- NODAGA-exendin-4 SUV; BCM_CLEAR: biomarker of beta cell mass computed with ^68^Ga- NODAGA-exendin-4 clearance.

There was a strong positive correlation between the biomarker of beta cell functional mass, i.e. glucose sensitivity of the beta cells, and both biomarkers of BCM in the subjects with obesity (Figure 4).

**Figure 4.**
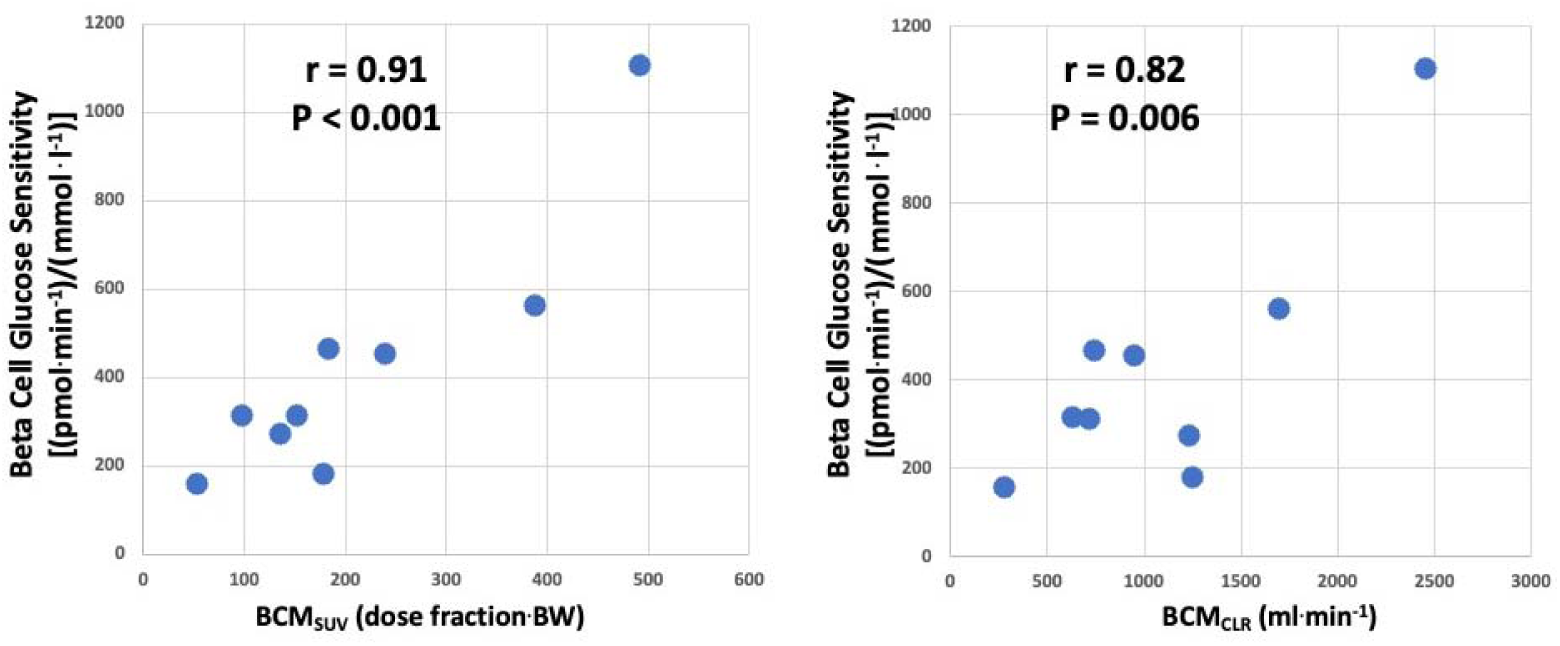
Left panel: relationship between the biomarker of BCM derived from ^68^Ga-NODAGA-exendin-4 SUV (BCM_SUV_; x axis) and the biomarker of beta cell functional mass derived from modeling glucose and hormone curves (beta cell glucose sensitivity; y axis) during a mixed meal tolerance test in people with obesity. Right panel: relationship between the biomarker of BCM derived from ^68^Ga-NODAGA-exendin-4 clearance (x axis) and the biomarker of beta cell functional mass derived from modeling glucose and hormone curves (beta cell glucose sensitivity; y axis) during a mixed meal tolerance test in people with obesity.

We could also derive biomarkers of average beta cell function (BCF) by dividing beta cell functional mass by either biomarker of BCM. In the subjects with obesity, BCF_SUV_ was 2.17±0.24 (range: 1.02 – 3.25), whereas BCF_CLEAR_ was 0.421±0.053 (range: 0.145 – 0.629).

## Discussion

This study employed a state-of-art assessment of beta cell functional mass and a modified ^68^Ga-NODAGA-exendin-4 PET-CT method to assess beta cell mass in participants with long standing type 1 diabetes and in individuals living with obesity. Two are the main findings of this study: 1. the biomarkers of BCM were 5- to 6.6-fold higher in subjects with obesity than in participants with type 1 diabetes, with an almost complete separation between the two groups (Figure 3a and 3b); 2. Both biomarkers of BCM were strongly related to beta cell functional mass in subjects with obesity (Figure 4).

Previous studies conducted with either different ligands (30)(31)(32) or with radioactive exendin-4 (33) have failed to provide the degree of separation we report in the present study between participants with type 1 diabetes and people without diabetes. Several reasons may be put forward to justify these differences, foremost of them: 1. The improvement in the specificity of ^68^Ga-exendin-4 pancreatic islet uptake, due to its correction with parotid glands as a reference tissue, and 2. the inclusion of pancreatic volume to compute a better estimate of total BCM.

As expected, the use of a reference region also mitigated the need of measuring arterial radioactivity curve to accurately assess ^68^Ga-exendin-4 uptake. The computation of specific pancreatic beta cell uptake taking into account abdominal aorta radioactivity curve resulted in virtually superimposable results, as already reported by others (22).

Pancreatic volume was found to be significantly lower in people with type 1 diabetes compared to nondiabetic subjects with obesity. This result was expected, because it was repeatedly reported in the past that pancreatic volume is decreased in individuals with type 1 diabetes (34), so much as to justify the tenet that type 1 diabetes is a disease which also affects the pancreatic exocrine tissue (35). However, it should be noted that the reduction in BCM reported in our study is a consequence not only of reduced pancreas mass, but also of reduced density of pancreatic uptake of ^68^Ga-NODAGA exendin-4, i.e. of reduced BCM per pancreatic volume unit. Thus, the findings of this study recapitulate the current knowledge regarding the components of BCM reduction in human type 1 diabetes.

The comparison between people with type 1 diabetes and people living with obesity highlights that in the latter the biomarker of BCM is 5-6 fold higher than in the former. This might be considered suboptimal when compared to the expected 10-20:1 ratio. However, to the best of our knowledge, this ratio appears to be by far the best one reported in previous papers of comparison of noninvasive biomarkers of BCM between T1DM individuals and people without diabetes (30)(15,31)(32)(33). Furthermore, the ratio herein reported is minimally dependent on the selection of a control group formed by people without diabetes living with obesity, because, in sharp contrast with data obtained in rodents, human obesity features a 20% increase in BCM (36). Moreover, as detailed in the Methods, we also did not include in our statistical analysis any of the findings of one OBESE outlier individual, in order to avoid inflating the estimates of the differences between the two groups and the statistical significance of the comparisons. The dynamic range of the biomarker of BCM both in the OBESE and in the participants with type 1 diabetes suggests that this tool may prove extremely useful in tracking in vivo the natural history of BCM in humans, even in people with type 1 diabetes.

We report that, in these subjects with obesity, there is a strong positive correlation between the biomarker of BCM and the biomarker of beta cell functional mass. Although this may be an expected finding, the degree of correlation may be stronger than it could be anticipated. To the best of our knowledge, there is no such previous assessment in vivo in humans. An early paper reported a coefficient of correlation of 0.42 between PET-CT scan derived biomarker of beta cell mass and beta cell functional mass as assessed by i.v. arginine stimulation superimposed to a hyperglycemic clamp (37). However, the specificity of [18F]fluoropropyl-(+)-dihydrotetrabenazine for pancreatic beta cells has been questioned (14), and the correlation was found pooling together people with normal glucose tolerance, or prediabetes or type 2 diabetes, but it was not evident in people with normal glucose tolerance only.

Our finding (Figure 4) may be explained in several, not mutually exclusive ways: 1. in people with obesity, there is, indeed, a very close relationship between BCM and beta cell functional mass, even within the normal glucose regulation; 2. a “winner’s curse” has overinflated the degree of correlation between BCM and beta cell functional mass; 3. it is a chance finding. The third possibility is dealt with by the relevantly low p values of the correlations (Figure 4). The first and second possibility represent a spectrum of the most plausible scenarios. Anyhow, our finding highlights that this technique can capture the relationship between BCM and beta cell functional mass, as long as the latter is investigated with state-of-art methods. On the other hand, studies with a much larger number of individuals with normal glucose regulation will be needed to provide accurate estimates of strength and shape (linear or curvilinear) of this relationship.

The combination between a biomarker of BCM and a biomarker of beta cell functional mass enabled us, for the first time, to assess a biomarker of bona fide beta cell “function”, i.e. the specific capability of beta cells to respond to a mixed meal. This biomarker also quantifies the “burden” posed on beta cells which compensate successfully for the metabolic stressors they are exposed to. It should be noted that the ranges of the biomarkers of BCM is almost 10-fold, whereas the ranges of the biomarkers of BCF is only 3- to 4-fold. This may be evidence that, at least in people with obesity, the “reserve” of BCF is unable to fully compensate for the lowest values of BCM, thereby unveiling an intrinsic “Achilles’ heel” of human beta cells.

In any case, this novel biomarker may enable one to gain more insight in vivo in humans regarding the complex processes underlying both compensation for and vulnerability to metabolic stressors, especially those which are risk factors for type 2 diabetes.

Our work is not devoid of limitations. First and foremost, the biomarker of BCM herein proposed, albeit a significant improvement on existing approaches, may be not absolutely specific in detecting beta cells, as suggested by the 5-6:1 ratio between people living with obesity and people with long-standing type 1 diabetes. On the other hand, the issue of the residual BCM, even in longstanding type 1 diabetes, is still open, and we presently cannot rule out the possibility that the residual signal detected in the participants with type 1 diabetes of the present study may reflect, at least in some of them, the presence of residual beta cells, “stunned” or “hiding” in a nonfunctional mode (38). Secondly, the number of subjects we studied is relatively small. However, the experimental design was rather demanding for the experimental subjects, because it required two long separate sessions on different days, with the use of rather sophisticated techniques of clinical physiology. Spreading our results, hopefully, will encourage other investigators to apply this method and rapidly increase the experience of the scientific community with it, thereby resulting in a fast appreciation of pros and cons of the present approach.

In summary, we report an improved ^68^Ga-exendin-4 method to estimate beta cell mass (BCM) in vivo in humans, with apparently a very promising potential to detect differences between different glucometabolic states. Furthermore, this approach has unveiled a very strong relationship between BCM and beta cell functional mass in people without diabetes living with obesity, to strongly suggest that in this population BCM plays a pivotal role in determining beta cell response to secretory stimuli. Altogether, these data imply that the noninvasive method herein reported is a novel, valuable adjunct in the study of the natural history of diabetes mellitus and related metabolic diseases in humans.

## Data Availability

All data produced in the present study are available upon reasonable request to the authors

## Acknowledgements

This work was supported by the Italian Ministry of University and Research (MUR) under the PRIN 2022 PNRR program – Project “In vivo separate assessment of pancreatic beta cell mass and beta cell function, from normal glucose regulation to longstanding type 2 diabetes mellitus, with the aid of a positron emission tomography-computed tomography (PET-CT) method” (Grant No. P2022445ES, CUP D53D23020950001), funded by the European Union – NextGenerationEU, Mission 4 “Education and Research”, Component 2 “From Research to Business”, Investment 1.1 and by the University of Parma through “Bando FIL – Quota incentivante 2019” to RCB and through “FIL” funds for research to ADC and RCB.

