## supplementary material for "Detection of pancreatic beta cell mass in vivo in humans: studies in individuals with long-standing type 1 diabetes and in individuals with obesity"

[1]Department of Medicine and Surgery, University of Parma, Italy; [2]Division of Endocrinology and Metabolic Diseases, Azienda Ospedaliera Universitaria Parma, Italy; [3]Division of Radiological Sciences, Azienda Ospedaliera Universitaria Parma, Italy; [4]Division of Nuclear Medicine, Azienda Ospedaliera Universitaria Parma, Italy; [5]Department of Medicine, University of Verona, Italy; [6]Division of Endocrinology, Diabetology and Metabolic Diseases, Azienda Ospedaliera Universitaria Integrata Verona, Italy

[†] These Authors jointly supervised this research

Please, address the correspondence to:
Riccardo C. Bonadonna
Division of Endocrinology, Diabetology and Metabolic Diseases
Ospedale Maggiore Borgo Trento
Piazzale Stefani 1
37126 Verona
Italy


### SUPPLEMENTARY MATERIAL

#### Mathematical Modeling of Pancreatic Beta Cell Secretory Response to a Mixed Meal Test

The analysis of the glucose and C-peptide curves during the mixed meal test in the present study is an evolution of the general strategy described in a previous publication with some modifications[1], which on turn was almost superimposable to the one introduced by other investigators[2,3]. At variance with the strategy applied by us in the past, we added also a component reflecting the incretin influence on mixed meal stimulated insulin secretion, as tracked through total GLP1 and total GIP concentration curves in plasma.

The kinetics of C-peptide is described with a two-compartment model, in which the two pools (1 and 2) exchange with each other and the irreversibile loss of the hormone is from pool 1, the same where C-peptide concentration is measured. C-peptide kinetic parameters are computed according to the equations by Van Cauter et al. [4].

Herein are the equations describing the model of glucose induced insulin secretion during an OGTT:

$$d\mathrm{cp}_1(\mathrm{t})/d\mathrm{t} = \mathrm{ISR(t)} + \mathrm{cp}_2 \cdot \mathrm{k}_{12} - (\mathrm{k}_{01} + \mathrm{k}_{21}) \cdot \mathrm{cp}_1 \qquad (1)$$

where ISR = insulin secretion rate, $cp_1$ = C-peptide mass in the sampling (accessibile) compartment, $cp_2$ = C-peptide mass in the remote compartment, $k_{12}$ and $k_{21}$ = rate constants of the exchange between the two C-peptide compartments, and $k_{01}$ = rate constant of the irreversibile loss of C-peptide from the accessibile compartment.

$$\mathrm{ISR(t)} = \mathrm{BSR} + \mathrm{SR^D(t)} + \mathrm{SR^P(t)} + \mathrm{SR^{INC}(t)} \qquad (2)$$

where BSR = basal insulin secretion rate, $SR^D$ = derivative or dynamic insulin secretion rate, $SR^P$ = proportional or static insulin secretion rate and $SR^{INC}(t)$ = incretin stimulated insulin secretion rate.

$$\mathrm{BSR} = \mathrm{CP_{ss}} \cdot \mathrm{V}_1 \cdot \mathrm{k}_{01} \qquad (3)$$

where $CP_{ss}$ is steady state unstimulated C-peptide concentration and $V_1$ is the volume of the accessibile compartment of C-peptide.

$$\mathrm{SR^D(t)} = \mathrm{X^D(t)} \cdot \tau^{-1} \qquad (4)$$

$$d\mathrm{X^D(t)} / d\mathrm{t} = \sigma^D \cdot \{[d\mathrm{G(t)}/d\mathrm{t}]/[\log(1.1+ \mathrm{t})]\} - \mathrm{X^D(t)} \cdot \tau^{-1} \qquad \text{if } d\mathrm{G(t)}/d\mathrm{t} > 0 \quad (5)$$

$$d\mathrm{X^D(t)} / d\mathrm{t} = - \mathrm{X^D(t)} \cdot \tau^{-1} \qquad \text{if } d\mathrm{G(t)}/d\mathrm{t} \leq 0 \quad (6)$$

where $\sigma^D$ = glucose sensitivity of derivative or dynamic insulin secretion, G = plasma glucose concentration, $X^D$ = C-peptide (insulin) mass made available for derivative/dynamic insulin secretion, $\tau$ = time constant of derivative/dynamic insulin secretion, and the term log(1.1 + t) accomodates the time-associated decline of $\sigma^D$ documented in humans during a hyperglycemic stimulus[5].

$$SR^P(t) = X^P(t) \cdot \delta^{-1} \qquad \text{if } X^P(t) > 0 \qquad (7)$$
$$SR^P(t) = 0 \qquad \text{if } X^P(t) \leq 0 \qquad (8)$$
$$dX^P(t) / dt = \sigma^P \cdot [G(t) - \theta] - X^P(t) \cdot \delta^{-1} \quad (9)$$

where $\sigma^P$ = glucose sensitivity of proportional/static insulin secretion, $X^P$ = C-peptide (insulin) mass made available for proportional/static insulin secretion, $\delta$ = time constant of proportional/static insulin secretion, $\theta$ = glucose threshold above which β-cell responds with proportional/static insulin secretion to plasma glucose concentration.

$$SR^{INC}(t) = X^{INC}(t) \cdot \gamma^{-1} \cdot \delta^{-1} \qquad (10)$$

In eq. 10, $X^{INC}(t)$ is the incretin-dependent mass of insulin which is made potentially available for secretion with the time constant $\gamma$. The fractional probability per time unit that $X^{INC}(t) \cdot \gamma^{-1}$ actually is released is equal to $\delta^{-1}$.

$$dX^P(t) / dt = \{\sigma^{GLP1} \cdot [GLP1(t) - GLP1bas] + \sigma^{GIP} \cdot [GIP(t) - GIPbas]\} - X^{INC}(t) \cdot \gamma^{-1}$$
$$\text{if } [GLP1(t) - GLP1bas] \text{ and } [GIP(t) - GIPbas] > 0 \ (11)$$

In eq. 11, GLP1(t) and GIP(t) are GLP1 and GIP concentrations, respectively, whereas GLP1bas and GIPbas are basal concentrations of GLP1 and GIP, respectively. Furthermore, $\sigma^{GLP1}$ and $\sigma^{GIP}$ are the sensitivities of beta cell to GLP1 and GIP action, respectively. Whenever [GLP1(t) – GLP1bas] or [GIP(t) – GIPbas] was ≤ 0, its value was set to 0.
Parameters were estimated by implementing this model of C-peptide secretion in the SAAM 1.2 software[6] (SAAM Institute, Seattle, WA). Numerical values of the unknown parameters were estimated by using nonlinear least squares. Weights were chosen optimally, i.e., equal to the inverse of the variance of the measurement errors, which were assumed to be additive, uncorrelated, with zero mean, and a constant coefficient of variation (CV) of 6%. In some studies, $\sigma^{GLP1}$ and/or $\sigma^{GIP}$ were estimated to be negligible.

The unknown parameters estimated by the model were:

1. $CP_{ss}$= steady state unstimulated C-peptide concentration (units: nmol/l);
2. $\sigma^D$ = glucose sensitivity of beta cell derivative/dynamic secretion (units: $[pmol \cdot (mmol \cdot l^{-1} \cdot min^{-1})^{-1}]$);
3. $\tau$ = time constant of beta cell derivative/dynamic secretion (units: min);
4. $\sigma^P$ = glucose sensitivity of beta proportional/static secretion (units: $[(pmol \cdot min^{-1}) \cdot (mmol \cdot l^{-1})^{-1}]$), which in the present paper is used as the biomarker of beta cell functional mass;
5. $\theta$ = glucose threshold of proportional/static secretion (units: mmol/l);
6. $\delta$ = time constant of proportional/static secretion (units: min)
7. $\sigma^{GLP1}$ = sensitivity of beta cell insulin to GLP1 (units: $[(pmol \cdot min^{-1}) \cdot (pmol \cdot l^{-1})^{-1}]$.
8. $\sigma^{GIP}$ = sensitivity of beta cell secretion to GIP (units: $[(pmol \cdot min^{-1}) \cdot (pmol \cdot l^{-1})^{-1}]$
9. $\gamma$ = time constant of provision of incretin stimulated insulin for insulin release.
